# Understanding Opportunities for Prescribing Pre-exposure Prophylaxis (PrEP) at Two Academic Medical Centers in a High Priority Jurisdiction for Ending the HIV Epidemic

**DOI:** 10.1101/2024.07.25.24310992

**Authors:** Moira C. McNulty, Katherine McGuckin, Eleanor E. Friedman, Matthew Caputo, Joseph A. Mason, Samantha A. Devlin, Mihai Giurcanu, Anu Hazra, Jessica P. Ridgway, Chad J. Achenbach

**Affiliations:** Department of Medicine, Section of Infectious Diseases, University of Chicago, Chicago, IL, USA; Havey Institute for Global Health, Feinberg School of Medicine, Northwestern University, Chicago, IL, USA; Department of Public Health Sciences, University of Chicago, Chicago, IL, USA; Department of Medicine, Division of Infectious Diseases, Feinberg School of Medicine, Northwestern University, Chicago, IL, USA

**Keywords:** HIV, pre-exposure prophylaxis, PrEP

## Abstract

**Introduction:** Pre-exposure prophylaxis (PrEP) is an effective, yet underutilized tool for HIV prevention. We sought to understand practice patterns and opportunities for prescribing PrEP across two large, urban, academic healthcare institutions in Chicago, Illinois.

**Methods:** We analyzed electronic medical record data from two institutions including encounters for persons ≥18 years of age with ≥1 negative HIV test between 1/1/2015-12/31/2021 who had indications for PrEP. Eligible encounters were those within a six-month window after STI diagnosis, or as long as injection drug use (IDU) was documented. We categorized encounters as inpatient, emergency department (ED), primary care, infectious disease (ID), obstetrics and gynecology/women’s health (OBGYN) and other outpatient settings. We performed bivariable and multivariable mixed effects regression models to examine associations, reporting odds ratios (or adjusted odds ratios) and 95% confidence intervals (OR, aOR, 95% CI).

**Results:** In total, 9644 persons contributed 53031 encounters that resulted in 4653 PrEP prescriptions. The two healthcare institutions had differing patient demographics; institution A had more 18–24 year-olds (58.3% vs 31.3%), more African Americans (83.8% vs 27.9%), and more women (65.7% vs 46.3%). Institution B had more White (40.6% vs 7.1%) and Hispanic persons (14.0% vs 4.2%), and more men who have sex with men (MSM) (15.2% vs 3.3%). Institution A had more eligible encounters in the ED (30.8% vs 7.3%) as well as in infectious disease, inpatient, OBYGN, and primary care settings. Institution B accounted for the majority of PrEP prescriptions (97.0%).

Adjusted models found lower odds of PrEP prescriptions in non-Hispanic Black (aOR 0.23 [0.16, 0.32]) and Latino (aOR 0.62 [0.44, 0.89]) patients, those with injection drug use (aOR 0.01 [0.00, 0.09]), men who have sex with women (aOR 0.36 [0.23, 0.56]), women who have sex with men (aOR 0.11 [0.06, 0.19]), and in the ED (ref) or OBGYN (0.11 [0.04, 0.27]) settings; while increased odds of PrEP prescription were associated with non-Hispanic White (ref) and MSM (aOR 24.87 [15.79, 39.15]) patients, and encounters at Institution B (aOR 1.78 [1.25, 2.53]) and in infectious disease (aOR [11.92 [7.65, 18.58]), primary care (aOR 2.76 [1.90, 4.01]), and other outpatient subspecialty settings (aOR 2.67 [1.84, 3.87]).

**Conclusions:** Institution A contained persons historically underrepresented in PrEP prescriptions, while institution B accounted for most PrEP prescriptions. Opportunities exist to improve equity in PrEP prescribing and across ED and OBGYN settings.

## Introduction

HIV pre-exposure prophylaxis (PrEP) is a key intervention for ending HIV transmission. The national Ending the HIV Epidemic (EHE) plan and many state plans call for increasing PrEP use in populations most impacted by HIV to reduce forward transmission (1–3). An estimated 1.1 million persons are considered at risk for HIV in the U.S. and may benefit from PrEP (4). PrEP is endorsed by the United States Preventive Services Task Force (USPSTF); criteria for eligibility and those who can most benefit from PrEP have been outlined by the Centers for Disease Control and Prevention (CDC) (5, 6). Although PrEP use has increased over the past several years, uptake is still below the estimated level needed to meet the goal of ending HIV transmission by 2030 (7–10). Furthermore, PrEP uptake remains variable across populations and geographic areas, often with the most vulnerable populations having the lowest PrEP uptake (7, 11, 12).

Within healthcare settings, many provider-level barriers to prescribing PrEP have been described, including lack of knowledge/awareness of who may benefit from PrEP on the parts of both provider and patient, lack of recognition of patient risk factors for HIV (i.e., sexual and social behaviors), and discomfort with prescribing PrEP (13–16). Barriers to prescribing PrEP and chronic under-prescribing for certain key populations have been documented; however, solutions for increasing appropriate PrEP prescribing at large, multispecialty academic medical centers have not been well described. Understanding healthcare systems-level trends in PrEP prescribing may offer insight into possible interventions to improve prescribing and uptake.

The objectives of this study were to 1) understand PrEP prescribing patterns and 2) opportunities for PrEP initiation at two academic medical centers within the greater Chicago area that serve populations in a high priority area for EHE.

## Methods

Data for this study included Electronic Data Warehouse (EDW) data from the University of Chicago Medicine (UCM) and Northwestern Medicine (NM) for adults (age 18 years or older) tested for HIV between January 1st, 2015 and December 31st, 2021. Patients were eligible for inclusion in the cohort if they had at least one negative HIV test during the study period, and did not receive a diagnosis of HIV, to ensure that only people eligible for PrEP were examined, mirroring the first step of the PrEP cascade. Data collected included demographic information, social history, medications, laboratory results for sexually transmitted infections (STI), ICD-10 codes for diagnosis of HIV, and encounter information.

We used the first negative HIV test as the entry date for the cohort, and we studied visit data (diagnoses, labs, etc.) after this entry date for PrEP indications. PrEP indications included one or more positive laboratory test results for chlamydia or gonorrhea or active syphilis as indicated by a rapid plasma reagin test ≥ 1:8. We also included anyone with injection drug use (IDU) noted in their social history. To mirror CDC clinical practice guidelines for PrEP, we created time periods to indicate when there was substantial risk of acquiring HIV infection (6). Medical encounters that occurred during these time periods were included in our assessment of missed opportunities for PrEP initiation. For persons with positive STI results, we examined encounters for six months after the test order. For IDU, we assumed continuous PrEP indication until documentation of cessation of IDU. To describe sexual behavior, we created sexual behavior categories using documented sex of the patient in combination with information on the sex of the partner and if the patient was currently sexually active. These categories included sexually active women who had sex with men (WSM), sexually active men who had sex with women (MSW), and sexually active men who had sex with men (MSM).

We removed medical encounters that we deemed unlikely to provide opportunities to offer PrEP. These encounters included medical procedures (imaging, dialysis, intravenous treatments) and encounters with non-prescribing providers (physical therapy/rehabilitation, audiology, speech pathology, behavioral health, nutrition). We categorized medical encounters to examine healthcare settings where PrEP was more or less likely to be prescribed. Categories included inpatient, emergency department (ED), primary care (including family medicine, primary care visits, urgent care settings, general pediatrics, and gerontology), infectious disease (ID), obstetrics and gynecology/women’s health (OBGYN/women’s health) and other outpatient settings (outpatient visits excluding OBGYN/women’s health, ID, or primary care).

PrEP prescriptions were determined by prescription for combination therapy with tenofovir disoproxil fumarate/emtricitabine, tenofovir alafenamide/emtricitabine, or the ingredients prescribed concurrently (emtricitabine and tenofovir disoproxil fumarate or tenofovir alafenamide) without the addition of other antiretroviral therapy (ART). If other ART prescriptions were present, we assumed that the patient was on either post-exposure prophylaxis (PEP) or treatment for HIV and did not count these persons among those on PrEP. To assign PrEP prescriptions to specific medical encounters and healthcare settings, we included all visits and locations two weeks prior to the order date of the PrEP prescription.

As data were obtained by two different healthcare institutions, research team consensus and standardization were sought before creation of categories or combining data to ensure as much continuity across variables as possible. Demographic information included race, ethnicity, age and electronic medical record (EMR)-reported sex. Age was categorized as follows: under 18 years, 18-24 years, 25-34 years, 35-44 years, 45-54 years, and 55 and older. Race and ethnicity information were combined to create the following categories: non-Hispanic Black, non-Hispanic white, Hispanic or Latino, Asian/Mideast Indian, more than one race, other, and unknown.

Descriptive statistics were performed using frequencies and percentages. Differences in characteristics between persons prescribed PrEP and persons not prescribed PrEP were compared using chi-square tests or Fisher’s exact tests. Bivariable and multivariable mixed effects regression models were created to examine associations, reporting odds ratios (or adjusted odds ratios) and 95% confidence intervals (OR, aOR, 95% CI). Mixed effect models were used to allow for multiple encounters by the same participant, as well as allow changing behavior over time for IDU, sexual activity, and sexual behavior. Two adjusted models were created to accommodate collinearity between documented sex and sexual behavior. All analyses included a variable to denote the differences between the two institutions while still providing a larger overview of the PrEP landscape in Chicago. For all models, complete case analysis was used, and if there were categories that did not vary by outcome (i.e., no PrEP prescriptions), this category was dropped from the model (i.e., as occurred when modeling missing age and missing sex). Data analyses were done in SAS (version 9.4, Cary North Carolina) and R (version 4.0.3; R Core Team). This study was approved by the Institutional Review Boards at the University of Chicago (IRB22-0237) and Northwestern University (STU00202938).

## Results

In total, 9,664 persons had both one or more negative HIV tests and one or more PrEP indications during the seven-year study period. These participants contributed 53,031 medical encounters to this analysis. The number of people with PrEP indications was close to equal between both institutions, with 46.6% from institution A and 53.4% from institution B. The two healthcare institutions had different patient demographic and risk factor distributions, with A having a larger number and proportion of 18-24 year olds (58.3% vs 31.3%), more African Americans (83.8% vs 27.9%), and more women (65.7% vs 46.3%). Institution A had more women reporting sexual activity with men (33.9% vs 23.0%) and more persons diagnosed with gonorrhea (35.8% vs 27.2%). While institution B had a larger number and greater proportion of White (40.6% vs 7.1%) and Hispanic persons (14.0% vs 4.2%), more men (50.5% vs 34.3%), and more men reporting sexual activity both with men and with women (MSM: 16.7% vs 3.9%, MSW: 25.5% vs 14.9%). Institution B also had more persons with a history of IDU (2.7% vs 0.6%), and more persons with active syphilis infection (11.1% vs 8.9%). Institutions A and B had similar proportions of people with positive chlamydia test results (72.5% vs 72.8%) (Table 1a).

**Table 1a:**
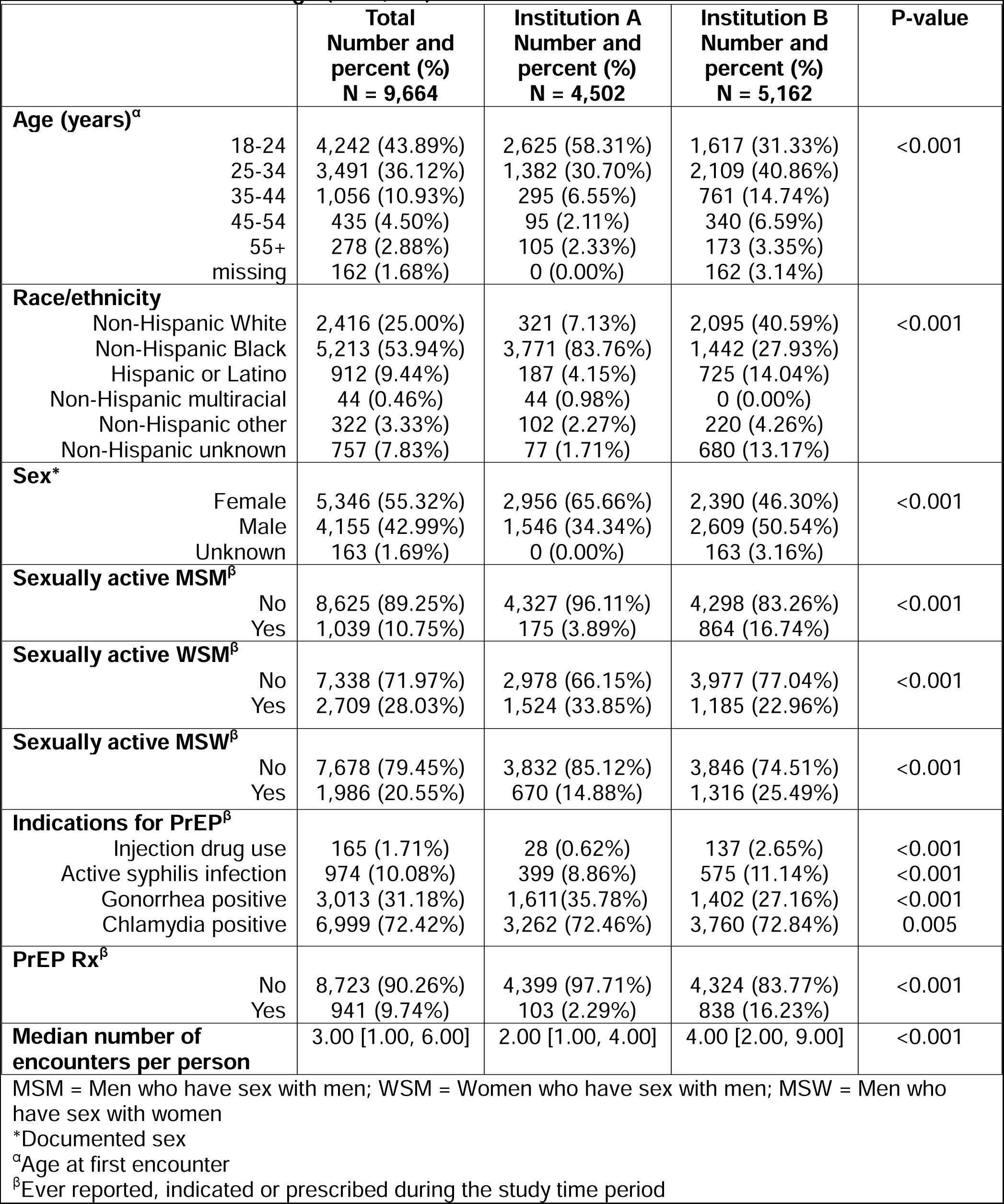
Characteristics of patients without HIV and indications for PrEP from 2015-2021 at two institutions in Chicago (n = 9,644)

|  | Total<br>Number and<br>percent (%)<br>N = 9,664 | Institution A<br>Number and<br>percent (%)<br>N = 4,502 | Institution B<br>Number and<br>percent (%)<br>N = 5,162 | P-value |
| --- | --- | --- | --- | --- |
| **Age (years)<sup>a</sup>** |  |  |  |  |
| 18-24 | 4,242 (43.89%) | 2,625 (58.31%) | 1,617 (31.33%) | <0.001 |
| 25-34 | 3,491 (36.12%) | 1,382 (30.70%) | 2,109 (40.86%) |  |
| 35-44 | 1,056 (10.93%) | 295 (6.55%) | 761 (14.74%) |  |
| 45-54 | 435 (4.50%) | 95 (2.11%) | 340 (6.59%) |  |
| 55+ | 278 (2.88%) | 105 (2.33%) | 173 (3.35%) |  |
| missing | 162 (1.68%) | 0 (0.00%) | 162 (3.14%) |  |
| **Race/ethnicity** |  |  |  |  |
| Non-Hispanic White | 2,416 (25.00%) | 321 (7.13%) | 2,095 (40.59%) | <0.001 |
| Non-Hispanic Black | 5,213 (53.94%) | 3,771 (83.76%) | 1,442 (27.93%) |  |
| Hispanic or Latino | 912 (9.44%) | 187 (4.15%) | 725 (14.04%) |  |
| Non-Hispanic multiracial | 44 (0.46%) | 44 (0.98%) | 0 (0.00%) |  |
| Non-Hispanic other | 322 (3.33%) | 102 (2.27%) | 220 (4.26%) |  |
| Non-Hispanic unknown | 757 (7.83%) | 77 (1.71%) | 680 (13.17%) |  |
| **Sex<sup>\*</sup>** |  |  |  |  |
| Female | 5,346 (55.32%) | 2,956 (65.66%) | 2,390 (46.30%) | <0.001 |
| Male | 4,155 (42.99%) | 1,546 (34.34%) | 2,609 (50.54%) |  |
| Unknown | 163 (1.69%) | 0 (0.00%) | 163 (3.16%) |  |
| **Sexually active MSM<sup>β</sup>** |  |  |  |  |
| No | 8,625 (89.25%) | 4,327 (96.11%) | 4,298 (83.26%) | <0.001 |
| Yes | 1,039 (10.75%) | 175 (3.89%) | 864 (16.74%) |  |
| **Sexually active WSM<sup>β</sup>** |  |  |  |  |
| No | 7,338 (71.97%) | 2,978 (66.15%) | 3,977 (77.04%) | <0.001 |
| Yes | 2,709 (28.03%) | 1,524 (33.85%) | 1,185 (22.96%) |  |
| **Sexually active MSW<sup>β</sup>** |  |  |  |  |
| No | 7,678 (79.45%) | 3,832 (85.12%) | 3,846 (74.51%) | <0.001 |
| Yes | 1,986 (20.55%) | 670 (14.88%) | 1,316 (25.49%) |  |
| **Indications for PrEP<sup>β</sup>** |  |  |  |  |
| Injection drug use | 165 (1.71%) | 28 (0.62%) | 137 (2.65%) | <0.001 |
| Active syphilis infection | 974 (10.08%) | 399 (8.86%) | 575 (11.14%) | <0.001 |
| Gonorrhea positive | 3,013 (31.18%) | 1,611 (35.78%) | 1,402 (27.16%) | <0.001 |
| Chlamydia positive | 6,999 (72.42%) | 3,262 (72.46%) | 3,760 (72.84%) | 0.005 |
| **PrEP Rx<sup>β</sup>** |  |  |  |  |
| No | 8,723 (90.26%) | 4,399 (97.71%) | 4,324 (83.77%) | <0.001 |
| Yes | 941 (9.74%) | 103 (2.29%) | 838 (16.23%) |  |
| **Median number of encounters per person** | 3.00 [1.00, 6.00] | 2.00 [1.00, 4.00] | 4.00 [2.00, 9.00] | <0.001 |

MSM = Men who have sex with men; WSM = Women who have sex with men; MSW = Men who have sex with women
\*Documented sex
<sup>a</sup>Age at first encounter
<sup>β</sup>Ever reported, indicated or prescribed during the study time period

The majority of encounters took place at institution B (36,761/53,031 (69.3%)), which is reflected in the median visit count for patients being significantly higher at institution B as compared to institution A (Median 4.00 interquartile range (IQR) [2.00-9.00] vs Median 2.00 IQR [1.00-4.00], p<0.001) (Table 1a, Table 1b).

**Table 1b.** Characteristics of encounters for patients without HIV and indications for PrEP from 2015-2021 at two institutions in Chicago (n=53,031)

|  | **Total**<br>**Number and**<br>**percent (%)**<br>**N=53,031** | **Institution A**<br>**Number and**<br>**percent (%)**<br>**N=16,270** | **Institution B**<br>**Number and**<br>**percent (%)**<br>**N=36,761** | **P-value** |
| --- | --- | --- | --- | --- |
| **Site of care** |  |  |  |  |
| Emergency department | 7,675 (14.47%) | 5,008 (30.78%) | 2,667 (7.25%) | <0.001 |
| Infectious disease | 1,158 (2.18%) | 514 (3.16%) | 644 (1.75%) |  |
| Inpatient | 1,107 (2.09%) | 680 (4.18%) | 427 (1.16%) |  |
| OBGYN/ women's health | 13,540 (25.53%) | 5,518 (33.92%) | 8,022 (21.82%) |  |
| Other Outpatient | 17,471 (32.94%) | 3,035 (18.65%) | 14,436 (39.27%) |  |
| Primary Care | 12,080 (22.78%) | 1,515 (9.31%) | 10,565 (28.74%) |  |
| **Sexually active MSM** |  |  |  | <0.001 |
| No | 43989 (82.95%) | 15660 (96.25%) | 28329 (77.06%) |  |
| Yes | 9042 (17.05%) | 610 (3.75%) | 8432 (22.94%) |  |
| **Sexually active WSM** |  |  |  | <0.001 |
| No | 37539 (70.79%) | 9477 (58.25%) | 28062 (76.34%) |  |
| Yes | 15492 (29.21%) | 6793 (41.75%) | 8699 (23.66%) |  |
| **Sexually active MSW** |  |  |  | <0.001 |
| No | 40157 (75.72%) | 14234 (87.49%) | 25923 (70.52%) |  |
| Yes | 12874 (24.28%) | 2036 (12.51%) | 10838 (29.48%) |  |
| **Indications for PrEP** |  |  |  |  |
| Intravenous drug use | 2362 (4.45%) | 379 (2.33%) | 1983 (5.39%) | <0.001 |
| Active syphilis infection | 6171 (11.64%) | 1646 (10.12%) | 4525 (12.31%) | <0.001 |
| Gonorrhea positive | 14559 (27.45%) | 4608 (28.32%) | 9951 (27.07%) | 0.003 |
| Chlamydia positive | 34539 (65.13%) | 11419 (70.18%) | 23120 (62.89%) | <0.001 |
| **PrEP Rx** |  |  |  | <0.001 |
| No | 48378 (91.23%) | 16131 (99.15%) | 32247 (87.72%) |  |
| Yes | 4653 (8.77%) | 139 (0.85%) | 4514 (12.28%) |  |

MSM = Men who have sex with men; WSM = Women who have sex with men; MSW = Men who have sex with women

The most frequent settings for total eligible encounters were other outpatient settings (17,471/53,031 (32.9%)), followed by OBGYN/women’s health (13,540/53,031 (25.5%)), primary care (12,080/53,031 (22. 8%)), and the ED (7,675/53,031 (14.5%)). Institution A contributed more total encounters (5,008/7,675 (65.3%)) to emergency visits as well as inpatient (680/1,107 (61.4%)) visits (Table 1b). Institution B contributed more total encounters to other outpatient encounters (14,436/17,471(82.6%)), ID (644/1,158 (55.6%)), OBGYN/women’s health (8,022/13,540 (59.2%)) and primary care (10,565/12,080 (87.5%)) (Table 1b).

Most encounters were deemed opportunities for PrEP initiation due to being six months after a chlamydia infection (34,539/53,031 (65.1%)), followed by encounters six months after a gonorrhea infection (14,559/53,031 (27.5%)), after a syphilis infection (6,171/53,031 (11.6%)), and encounters in which IDU was documented (2,362/53,031 (4.5%)). Overall, 941 total people received PrEP prescriptions, with the majority attending institution B (89.1%). These prescriptions were within a two-week window of 4,653 encounters (4,653/53,031 (8.8%)).

When examining unadjusted associations between missed opportunities for PrEP and various characteristics, many factors were significantly associated with a PrEP prescription (Table 2). Attending institution B resulted in nearly six times the odds of PrEP prescription (OR 5.75 95%CI (3.39, 9.76)) than attending institution A. Being Black, Hispanic or of unknown race resulted in fewer PrEP prescriptions than being White, ranging from being 0.15 times as likely to being 0.47 times as likely to obtain a PrEP prescription (Black: OR 0.15 95%CI (0.10, 0.22), Hispanic: OR 0.36 95%CI (0.21, 0.64), and unknown race/ethnicity OR 0.47 95% CI (0.26, 0.86)). Being older than 24 years of age was associated with increased odds of PrEP prescription compared to individuals ages 18-24, ranging from 3.23 to 5.20 times as likely (25-34 years: OR 3.23 95%CI (2.24, 4.66), 35-44 years: OR 5.20 95%CI (3.43, 7.88), 45-54 years: OR 4.53 95%CI (2.73, 7.52) and 55+ years: OR 3.53 95%CI (1.84, 6.80)). Males had nearly 66 times the odds of obtaining a PrEP prescription as females (OR 63.16 95%CI (34.32, 116.23)).

**Table 2:**
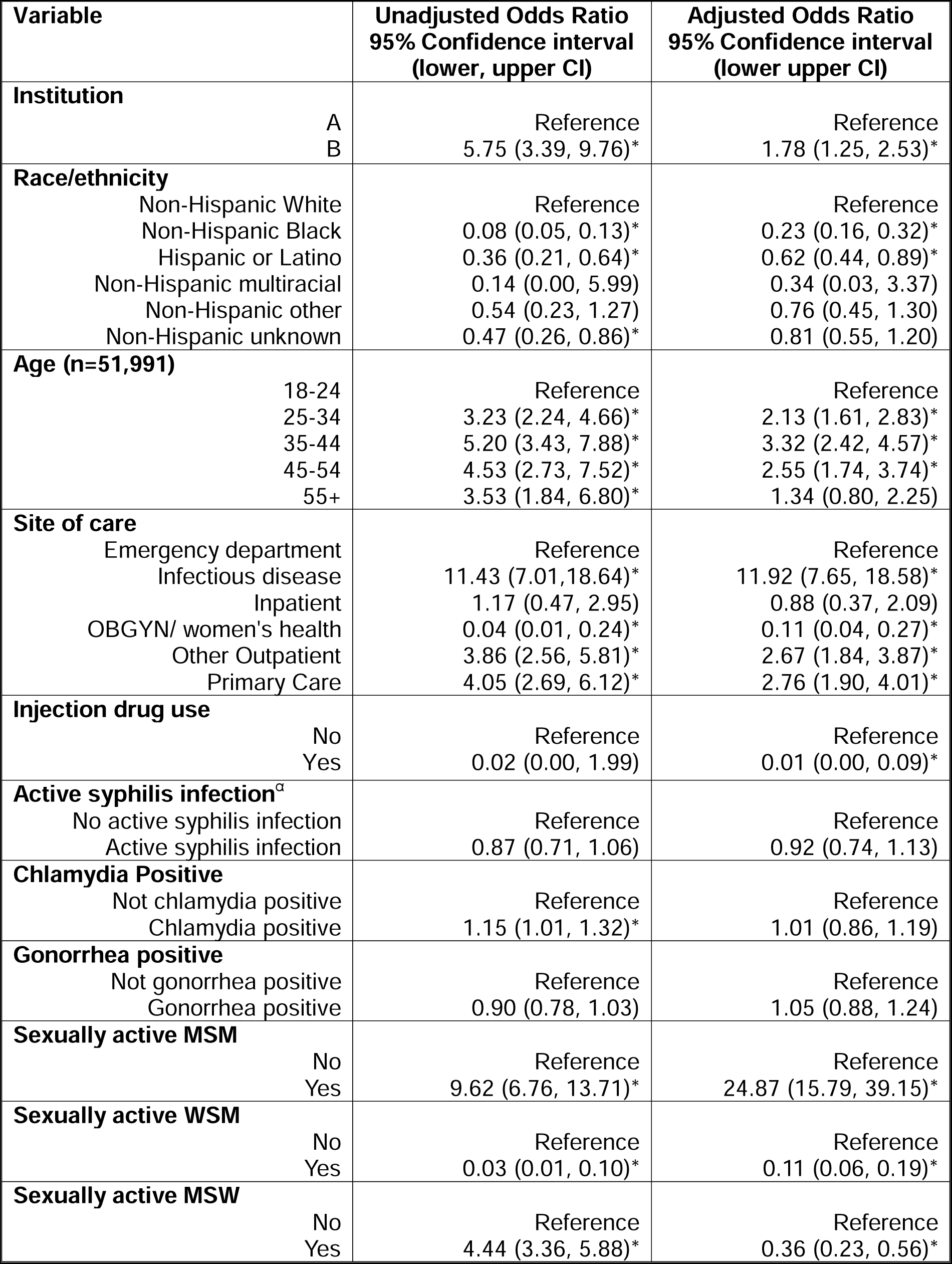
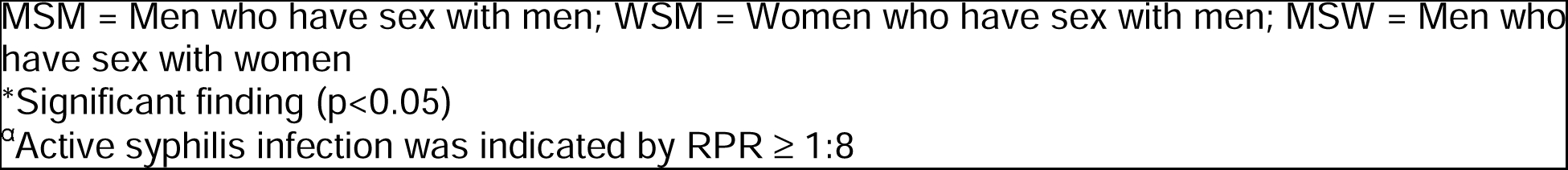
Odds ratios and confidence intervals for mixed effects models examining PrEP prescriptions at the encounter level for a model containing sexual behavior (n=53,031)

| Variable | Unadjusted Odds Ratio<br>95% Confidence interval<br>(lower, upper CI) | Adjusted Odds Ratio<br>95% Confidence interval<br>(lower upper CI) |
| --- | --- | --- |
| **Institution** |  |  |
| A | Reference | Reference |
| B | 5.75 (3.39, 9.76)* | 1.78 (1.25, 2.53)* |
| **Race/ethnicity** |  |  |
| Non-Hispanic White | Reference | Reference |
| Non-Hispanic Black | 0.08 (0.05, 0.13)* | 0.23 (0.16, 0.32)* |
| Hispanic or Latino | 0.36 (0.21, 0.64)* | 0.62 (0.44, 0.89)* |
| Non-Hispanic multiracial | 0.14 (0.00, 5.99) | 0.34 (0.03, 3.37) |
| Non-Hispanic other | 0.54 (0.23, 1.27) | 0.76 (0.45, 1.30) |
| Non-Hispanic unknown | 0.47 (0.26, 0.86)* | 0.81 (0.55, 1.20) |
| **Age (n=51,991)** |  |  |
| 18-24 | Reference | Reference |
| 25-34 | 3.23 (2.24, 4.66)* | 2.13 (1.61, 2.83)* |
| 35-44 | 5.20 (3.43, 7.88)* | 3.32 (2.42, 4.57)* |
| 45-54 | 4.53 (2.73, 7.52)* | 2.55 (1.74, 3.74)* |
| 55+ | 3.53 (1.84, 6.80)* | 1.34 (0.80, 2.25) |
| **Site of care** |  |  |
| Emergency department | Reference | Reference |
| Infectious disease | 11.43 (7.01, 18.64)* | 11.92 (7.65, 18.58)* |
| Inpatient | 1.17 (0.47, 2.95) | 0.88 (0.37, 2.09) |
| OBGYN/ women's health | 0.04 (0.01, 0.24)* | 0.11 (0.04, 0.27)* |
| Other Outpatient | 3.86 (2.56, 5.81)* | 2.67 (1.84, 3.87)* |
| Primary Care | 4.05 (2.69, 6.12)* | 2.76 (1.90, 4.01)* |
| **Injection drug use** |  |  |
| No | Reference | Reference |
| Yes | 0.02 (0.00, 1.99) | 0.01 (0.00, 0.09)* |
| **Active syphilis infection<sup>a</sup>** |  |  |
| No active syphilis infection | Reference | Reference |
| Active syphilis infection | 0.87 (0.71, 1.06) | 0.92 (0.74, 1.13) |
| **Chlamydia Positive** |  |  |
| Not chlamydia positive | Reference | Reference |
| Chlamydia positive | 1.15 (1.01, 1.32)* | 1.01 (0.86, 1.19) |
| **Gonorrhea positive** |  |  |
| Not gonorrhea positive | Reference | Reference |
| Gonorrhea positive | 0.90 (0.78, 1.03) | 1.05 (0.88, 1.24) |
| **Sexually active MSM** |  |  |
| No | Reference | Reference |
| Yes | 9.62 (6.76, 13.71)* | 24.87 (15.79, 39.15)* |
| **Sexually active WSM** |  |  |
| No | Reference | Reference |
| Yes | 0.03 (0.01, 0.10)* | 0.11 (0.06, 0.19)* |
| **Sexually active MSW** |  |  |
| No | Reference | Reference |
| Yes | 4.44 (3.36, 5.88)* | 0.36 (0.23, 0.56)* |

MSM = Men who have sex with men; WSM = Women who have sex with men; MSW = Men who have sex with women
\*Significant finding ( $p < 0.05$ )
<sup>a</sup>Active syphilis infection was indicated by $RPR \geq 1:8$

Having appointments in the ID department, primary care, or other outpatient setting all increased the odds of obtaining a PrEP prescription as compared to an appointment in the ED (ID: OR 11.43 95% CI (7.01, 18.64), other outpatient OR 3.86 95%CI (2.56, 5.81), primary care OR 4.05 95%CI (2.69, 6.12)). However, having an encounter in the OBGYN/women’s health department was associated with decreased odds of PrEP prescription (OR 0.04 95%CI (0.01, 0.24)). In terms of PrEP indications, only recent infection with chlamydia was significantly associated with PrEP prescription (OR 1.15 95%CI (1.01, 1.32)). Identifying as a male who had sexual partners with either men or women was associated with increased odds of PrEP prescription of 4 or 10 times, respectively, that of those not reporting these behaviors (MSM: OR 9.62 95%CI (6.76, 13.71), MSW: OR 4.44 95%CI (3.36, 5.88)). However, women who had sex with men had lower odds of PrEP prescriptions compared to those not reporting this behavior (WSM: OR 0.03 95%CI (0.01, 0.10)).

When examining adjusted associations in a model including sexual behavior but not sex, results were largely similar to those seen in unadjusted analyses, although many of the magnitude of point estimates and confidence intervals were attenuated (Table 2). A few results were no longer significant including the odds of PrEP prescription for those 55 years and older (aOR 1.34 95%CI (0.80, 2.25)) and those who received a recent positive test result for chlamydia (aOR 1.01 95%CI (0.86, 1.19)). However, the association of sexually active MSM having a higher odds of PrEP prescription increased to 24.87 (95%CI (15.79, 39.15)) from 9.62 (95%CI (6.76, 13.71)). IDU increased in magnitude of association in the multivariate model with reduced odds of PrEP prescription for those reporting this behavior compared to those who did not report this behavior (aOR 0.01 95%CI (0.00, 0.09)). The association for MSW in the sexual behavior adjusted model reversed direction, decreasing from 4.44 times to 0.36 times the odds of PrEP prescription (95%CI (0.23, 0.56)) (Table 2).

When examining the adjusted model including sex, results were largely similar to those obtained when modeling sexual behavior (Supplemental Table 1). Adjusted models containing sex resulted in increased odds for male sex, though slightly attenuated from unadjusted results (aOR 32.36, 95%CI (21.07, 49.70)).

## Discussion

Underutilization of PrEP can only improve if we understand prescribing patterns and opportunities to offer PrEP among patients with clear indications. In this study, we examined PrEP prescribing at two large academic healthcare systems in Chicago and found opportunities to improve PrEP delivery and awareness in settings where patients are accessing sexual health services, including the ED and OBGYN/women’s health clinics. Overall, approximately 10% of the patients who were included within our analysis received a prescription for PrEP. This prescription rate is slightly higher than studies within similar settings that have found PrEP prescription rates between 1% and 8% (17–19).

We found that persons who identified as Black or Hispanic and younger individuals were less likely to be prescribed PrEP. This may be related to less PrEP prescribing overall at Institution A, where approximately 84% of patients with indications for PrEP were Black and 58% in the 18–24-year-old age category. However, this pattern has been reported in other studies. Indeed, older age has been correlated with increased odds of PrEP prescription (18, 20), while Pitts et al. found that White and Hispanic patients were more likely than Black patients to receive PrEP at an urban medical center (21). In a study that examined individuals who tested positive for an STI within the ED, Black patients were less likely to receive follow-up care (20). However, Agovi et al. found that among individuals who were prescribed PrEP within an urban safety-net health system, the majority were racial or ethnic minorities (18).

In addition to racial and age disparities, we also found that males had significantly higher odds of receiving a PrEP prescription than females. When examining this in the adjusted models, this result was largely accounted for by prescribing for MSM. Yumori et al. investigated missed opportunities for PrEP among individuals testing positive for an STI at a large academic medical center and found that both multi-site testing for gonorrhea/chlamydia and syphilis testing were more likely to be performed among men compared to women; furthermore, women were more likely to be inadequately screened for HIV, and PrEP was discussed almost exclusively with men (22). In a comparable analysis of patients who were seen at an urban municipal medical center, despite accounting for over half of the patients with an indication for PrEP, no women received a PrEP prescription (21). Within these studies, men typically have “high risk sexual behavior” as their primary indication for PrEP (17, 18, 23); however, the most common indication for PrEP among women is having a sexual partner living with HIV (23, 24). There is a clear need for comprehensive documentation within the EMR of risk factors (i.e., information regarding sexual partners) that are more pertinent to women if this gap in HIV prevention is to be addressed adequately (25).

In large healthcare systems with acute ED- and hospital-based care, as well as primary and subspecialty care, targeted efforts to increase PrEP awareness and uptake in areas where indicated patients are accessing care may be more effective than a systems-wide initiative. In this study, PrEP prescribing was associated with encounters in the ID department, primary care (23), and other subspecialty clinics, though many encounters for patients with PrEP indications also occurred in the ED and OBGYN/women’s health, where we found reduced odds of PrEP prescriptions. In the ED, collecting information on sexual behavior and STIs may facilitate identifying patients who may benefit from PrEP. In the OBGYN/women’s health setting, patients are overwhelmingly heterosexual women who are often not considered to be at risk for HIV acquisition, yet within certain populations in Chicago women account for up to 30% of new HIV diagnoses, much higher than the national average (26, 27). Further, clinician confidence with discussing or initiating PrEP may be a barrier, particularly in the ED where continuity of follow up for PrEP monitoring may not be clearly established and more acute medical concerns may be prioritized.

Our results can guide interventions or health system policy changes, yet additional information on barriers to prescribing should be considered. Qualitative studies have identified barriers to prescribing PrEP, including insufficient provider training, competing priorities and limited time during appointments, side effect concerns, and health care provider workload (28). In addition to developing EMR alerts and making consultation available, provider training, protocols for PrEP prescribing, and clinical decision support tools will be important strategies, particularly for clinicians not accustomed to prescribing PrEP. However, in a large trial of a clinical decision support tool that alerted providers of patient indications using an EMR-based algorithm, no differences in prescribing were seen among clinicians who don’t treat HIV, suggesting high barriers to prescribing PrEP (29). Even so, accurate algorithms may rely on documentation of sexual behaviors, which we found likely to be under-documented for at least one participating institution. Alternative models can be considered, such as same-day sexual health appointments for those presenting with sexual health or STI-related concerns, including to the ED (15, 29–31). Other authors have suggested methods to streamline PrEP prescribing such as PrEP navigation, same-day initiation, and easing the complexity of PrEP care that deters many from prescribing (29, 31).

There were several limitations to this study. Some information, including insurance data, were unavailable from both healthcare systems, and thus could not be included in the combined analysis. Social, sexual, and drug use behaviors may not have been consistently or accurately collected within structured fields within the EMR; thus, it was possible that indications for PrEP were misclassified. Interpretation of syphilis test results can be complicated and at times difficult to determine an active versus past infection. The RPR cutoff of 1:8 or greater may have excluded some patients who had indications for PrEP; a more nuanced analysis of syphilis indicators for PrEP would be informative. Using all encounters within a two-week window of PrEP prescriptions may have overestimated the actual number of departments/locations that prescribed PrEP. However, our approach did consider timing from prescription order to patient pickup, as well as for delays from insurance prescription drug coverage. Finally, it is unknown how many patients declined PrEP when offered it or how many patients received education about PrEP but not a prescription. Strengths of our study include the use of data from two different healthcare systems in Chicago and the length of the study period.

## Conclusions

In this multi-institution study of PrEP prescribing, we found opportunities to improve PrEP prescribing where patients are accessing care for sexual health, as well as opportunities to improve equitable PrEP prescribing among young Black and Latinx populations highly impacted by HIV. Our study results can inform targeted interventions at the healthcare systems level to improve PrEP prescribing and uptake for indicated patients.

## Data Availability

All data produced in the present work are contained in the manuscript.

## Funding & Acknowledgments

This publication was made possible with support from the Third Coast Center for AIDS Research (CFAR), an NIH funded center (P30 AI117943). Moira McNulty has served on an advisory board for Gilead Sciences. Jessica Ridgway has received consulting fees from Gilead Sciences. Anu Hazra receives research support from Gilead Sciences, and has served on advisory boards for Gilead Sciences and Viiv Healthcare. All other authors report no conflicts of interest. Eleanor Friedman has received funds as part of a Gilead FOCUS grant.

