## Supplemental Table 1 for "Understanding Opportunities for Prescribing Pre-exposure Prophylaxis (PrEP) at Two Academic Medical Centers in a High Priority Jurisdiction for Ending the HIV Epidemic"

**Supplemental Table 1: Odds ratios and confidence intervals for mixed effects models examining PrEP prescriptions at the encounter level for a model containing sex (n=53,031)**

| **Variable** | **Unadjusted Odds Ratio 95% Confidence interval (lower, upper CI)** | **Adjusted Odds Ratio 95% Confidence interval**  **(lower, upper CI)** |
| --- | --- | --- |
| **Institution**  A  B | Reference  5.75 (3.39, 9.76)* | Reference  2.48 (1.66, 3.68)* |
| **Race/ethnicity**  Non-Hispanic White  Non-Hispanic Black  Hispanic or Latino  Non-Hispanic multiracial  Non-Hispanic other  Non-Hispanic unknown | Reference  0.08 (0.05, 0.13)*  0.36 (0.21, 0.64)*  0.14 (0.00, 5.99)  0.54 (0.23, 1.27)  0.47 (0.26, 0.86)* | Reference  0.15 (0.10, 0.22)*  0.47 (0.31, 0.73)*  0.25 (0.02, 3.16)  0.64 (0.34, 1.21)  0.52 (0.33, 0.84)* |
| **Age (n=51,991)**  18-24  25-34  35-44  45-54  55+ | Reference  3.23 (2.24, 4.66)*  5.20 (3.43, 7.88)*  4.53 (2.73, 7.52)*  3.53 (1.84, 6.80)* | Reference  2.15 (1.57, 2.94)*  3.26 (2.29, 4.64)*  2.41 (1.58, 3.69)*  1.45 (0.82, 2.56) |
| **Sex**^α^ **(n=51,973)**  Female  Male | Reference  63.16 (34.32, 116.23)* | Reference  32.36 (21.07, 49.70)* |
| **Site of care**  Emergency department  Infectious disease  Inpatient  OBGYN/ women's health  Other Outpatient  Primary Care | Reference  11.43 (7.01,18.64)*  1.17 (0.47, 2.95)  0.04 (0.01, 0.24)*  3.86 (2.56, 5.81)*  4.05 (2.69, 6.12)* | Reference  9.16 (5.81, 14.43)*  0.98 (0.40, 2.43)  0.16 (0.05, 0.57)*  2.10 (1.44, 3.07)*  2.18 (1.49, 3.19)* |
| **Injection drug use**  No  Yes | Reference  0.02 (0.00, 1.99) | Reference  0.00 (0.00, 0.05)* |
| **Active syphilis infection**^β^  No active syphilis infection  Active syphilis infection | Reference  0.87 (0.71, 1.06) | Reference  0.84 (0.67, 1.04) |
| **Chlamydia Positive**  Not chlamydia positive  Chlamydia positive | Reference  1.15 (1.01, 1.32)* | Reference  1.09 (0.92, 1.28) |
| **Gonorrhea positive**  Not gonorrhea positive  Gonorrhea positive | Reference  0.90 (0.78, 1.03) | Reference  1.04 (0.88, 1.24) |

MSM = Men who have sex with men; WSM = Women who have sex with men; MSW = Men who have sex with women

*Significant finding (p<0.05)

^α^Documented sex

^β^Active syphilis infection was indicated by RPR ≥ 1:8
